# Sex Differences in the Acute Effects of Oral THC: A Randomized, Placebo-Controlled, Crossover Human Laboratory Study

**DOI:** 10.1101/2023.11.29.23299193

**Authors:** Ardavan Mohammad Aghaei, Lia Urban Spillane, Brian Pittman, L. Taylor Flynn, Joao P. De Aquino, Anahita Bassir Nia, Mohini Ranganathan

## Abstract

**Rationale:** Recent reports have shown increased cannabis use among women, leading to growing concerns about cannabis use disorder (CUD). Some evidence suggests a faster progression to addiction in women, known as the “telescoping effect.” While there is preclinical evidence suggesting biological sex influences cannabinoid effects, human research remains scant. We investigated sex differences in the response to oral tetrahydrocannabinol (THC) in humans.

**Methods:** 56 healthy men and women with prior exposure to cannabis but no history of CUD participated in a randomized, placebo-controlled, human laboratory study where they received a single 10 mg dose of oral THC (dronabinol). Subjective psychoactive effects were assessed by the visual analog scale of “high”, psychotomimetic effects by the Clinician-Administered Dissociative Symptoms Scale and Psychotomimetic States Inventory, verbal learning and memory by Rey Auditory Verbal Learning Test (RAVLT), and physiological effects by heart rate. Outcomes were regularly measured on the test day, except for the RAVLT, which was assessed once. Peak differences from baseline were analyzed using a nonparametric method for repeated measures.

**Results:** Oral THC demonstrated significant dose-related effects in psychotomimetic and physiological domains, but not in RAVLT outcomes. A notable interaction between THC dose and sex emerged concerning the subjective “high” scores, with women reporting heightened sensations (p=0.05). No other significant effects of sex and THC dose interaction were observed.

**Conclusion:** Oral THC yields similar psychotomimetic and physiological effects across sexes, but women may experience a pronounced subjective psychoactive effect. Further research is needed to identify individual vulnerabilities and facilitate tailored interventions addressing CUD.

## 1. Introduction

With the shifting landscape of cannabis legalization worldwide and especially in the U.S., both cannabis usage and its associated disorders are anticipated to surge (Weinberger et al. 2022). Over the last decade, cannabis consumption has consistently risen (Center for Behavioral Health Statistics and Quality 2023; Mitchell et al. 2020). The National Survey on Drug Use and Health (NDSU) reveals that in 2021, 18.7% of Americans (52.4 million) aged 12 or older used cannabis, with 0.9% (2.6 million) being first-time users. Alarmingly, 5.8% (16.2 million) met the criteria for cannabis use disorder (CUD) (Center for Behavioral Health Statistics and Quality 2023). As cannabis becomes more accessible, understanding the individual biopsychosocial vulnerabilities that lead to the development of CUD is imperative to develop preventive and treatment measures.

Over the past decade, the incidence of first-time cannabis use has risen among women, and the gap between the rates of cannabis use in men and women has narrowed (Chapman et al. 2017; Johnson et al. 2015). Epidemiologic data suggest that women develop CUD sooner than men following their primary exposure to cannabis use. This compressed timeline from first use to CUD is referred to as the “telescopic phenomenon” (Gräfe et al. 2023; Hernandez-Avila et al. 2004; Khan et al. 2013). Similarly, women are more likely to experience cannabis withdrawal symptoms and might experience them more severely (Copersino et al. 2010; Herrmann et al. 2015). In addition, women are more likely to report a greater impact of cannabis use on their quality of life (Lev-Ran et al. 2012).

Preclinical evidence suggests that sex can be a moderating factor of acute cannabis effects (Bassir Nia et al. 2018; Calakos et al. 2017; Cooper and Craft 2018). Human studies have investigated sex differences in the acute effects of cannabis as well as delta-9-tetrahydrocannabinol (THC), the active constituent of cannabis, using oral, Intravenous (IV), and inhaled routes with mixed results. For instance, some studies have reported no sex differences in the acute subjective effects of cannabis or THC (Cocchetto et al. 1981; Matheson et al. 2020; Mathew et al. 2003), or stronger effects in men (Haney 2007; Penetar et al. 2005), while other studies show a heightened sensitivity to subjective effects in women (Bassir Nia et al. 2022; Cooper and Haney 2009; 2014; Fogel et al. 2017; Makela et al. 2006). In particular, the literature with oral THC or THC-containing cannabis also has inconsistent results (Fogel et al. 2017; Haney 2007; MacNair et al. 2023; Sholler et al. 2021) **(Table S1)**. These may be related to the formulation of THC or cannabis, the latter while more generalizable, introduces confounds due to the presence of other compounds in herbal cannabis (MacNair et al. 2023; Sholler et al. 2021). Another important variable to consider in reviewing the literature is the inclusion of frequent users of cannabis, who could have different sensitivity to the dose-related effects of THC compared to infrequent users (Fogel et al. 2017).

We have previously examined sex differences in the acute effects of THC in a double-blind, placebo-controlled, randomized human laboratory study using a well-validated paradigm with IV THC (Carbuto et al. 2012; Englund et al. 2012). We demonstrated dose-related sex differences in the acute subjective effects such that women experienced a greater subjective “high” at the lower THC dose without experiencing greater psychotomimetic or cognitive effects (Bassir Nia et al. 2022). Importantly, IV THC enabled us to control dose delivery, variable absorption rates observed with oral or inhaled administration, and first-pass hepatic metabolism, but was limited in demonstrating ecological validity of the findings. Since cannabis is usually smoked or ingested, sex differences in the absorption rate, metabolism, and peak concentration of cannabinoids (Lunn et al. 2019; Nadulski et al. 2005; Narimatsu et al. 1991; Wall et al. 1983) could contribute to the overall sex differences in the cannabis effect we observe in the community. Studying the acute effects of oral THC can provide more generalizable findings compared to IV THC. It is important to note that data from oral administration should not be extrapolated to smoked or vaporized cannabinoids.

In this study, we sought to investigate the sex differences in acute subjective psychoactive, psychotomimetic, physiological, and cognitive (verbal learning and memory) effects following a single fixed dose of oral synthetic THC (dronabinol) in healthy individuals with prior exposure to cannabis but not meeting the criteria for CUD.

## 2. Methods

This randomized, placebo-controlled, double-blind, human laboratory study evaluated the acute subjective psychoactive, psychotomimetic, physiological effects and verbal learning and memory following a single fixed dose of oral THC in 56 healthy individuals (33 women and 23 men). The study protocol was approved by the Institutional Review Boards of the VA Connecticut Healthcare System and Yale University School of Medicine.

### 2.1 Participants

Healthy men and women between the ages of 18 and 55 years were recruited from the community using flyers, digital advertisements, and word-of-mouth. Inclusion criteria included having used cannabis at least once over the three months prior to study participation, without ever meeting the Diagnostic and Statistical Manual of Mental Disorders (DSM)-5 criteria for CUD. Exclusion criteria included lifetime or current DSM-5 major psychiatric disorders such as schizophrenia, schizoaffective disorder, or bipolar disorder; major or clinically unstable medical conditions; major current or recent stressors over the previous 6 weeks (based on clinical interview); IQ less than 80 per Wechsler Test of Adult Reading; or blood donation within the previous 8 weeks. Participants were also excluded if they met DSM-5 criteria for lifetime CUD and other substance use disorders (except for tobacco) within the last three months. Participants were required to have negative urine toxicology and pregnancy tests.

### 2.2 Screening

After an initial phone screening, participants were provided with a full, written explanation of the study procedures. All the procedures were explained verbally by trained staff, and participants completed a brief questionnaire to confirm their understanding of the risks of the study. Participants were provided enough time to ask questions, addressed by study physicians and research staff. Once signed informed consent was obtained, participants underwent medical and psychiatric examinations, and blood and urine samples were collected. The Structured Clinical Interview (SCID) for DSM-5 was conducted by trained staff. Participants’ sex refers to their self-designation of sex assigned at birth. For clarity and consistency in this manuscript, we refer to female participants as “women” and male participants as “men”. The participants were required to have negative urine toxicology. The absence of pregnancy in women was confirmed by urine pregnancy testing on screening and the morning of each test day (**Figure 1**).

**Figure 1:**
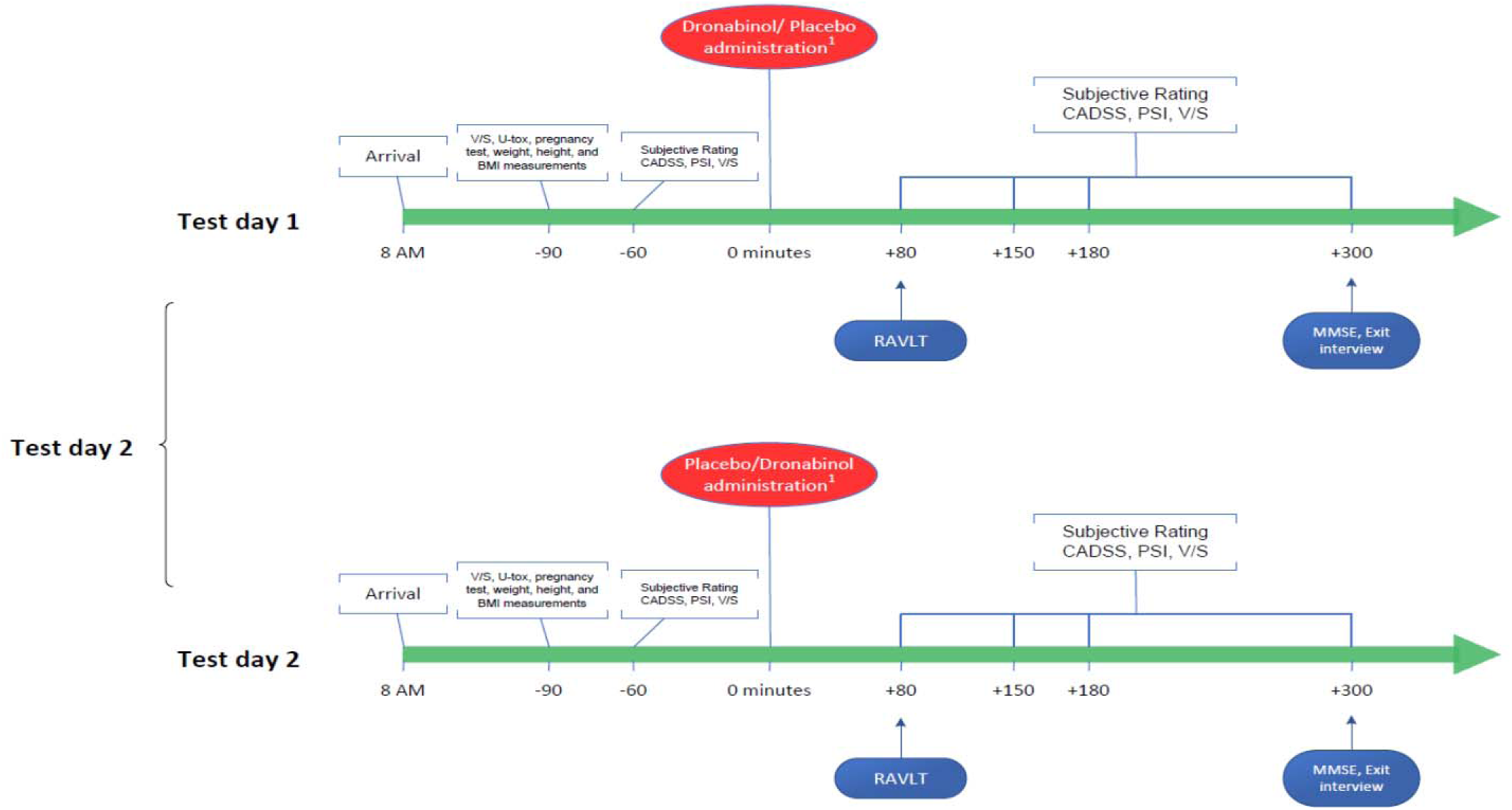
Study Design and schedule of procedures on each test day. abbreviations: BMI, Body Mass Index; CADSS, Clinician-Administered Dissociative States Scale; MMSE, Mini-Mental State Examination; PSI, Psychotomimetic States Inventory. ^1^ The number of patients receiving placebo and dronabinol in any order was counterbalanced.

### 2.3 Assessments

#### 2.3.1 Subjective psychoactive effects

The subjective “high” induced by THC was gauged using a visual analog scale (VAS). Participants were instructed to rate the intensity of their perceived “high” on a 100mm line, with reference points at 0 (indicating “not at all”) and 100 (indicating “extremely”). These assessments were conducted both before and at multiple intervals following THC/placebo administration. The efficacy of this measure in capturing THC effects has been validated in prior research (D’Souza et al. 2008).

#### 2.3.2 Psychomimetic and dissociative effects

To evaluate thought and perceptual changes, we employed the Clinician-Administered Dissociative Symptoms Scale (CADSS) and the Psychotomimetic States Inventory (PSI). Both the CADSS and the PSI are recognized for their validity and have demonstrated sensitivity to THC-induced effects (Bassir Nia et al. 2022). The CADSS is a 28-item questionnaire containing 23 participant-rated and five observer-rated items aimed to capture perceptual alterations and dissociative effects on a scale of 0 (not at all) to 4 (extreme) (Bremner et al. 1998). The PSI has 48 items, each scored on a Likert scale of 0 to 3, covering the six domains of psychotic-like experiences (Mason et al. 2008). A trained rater administered these behavioral ratings.

#### 2.3.3 Verbal Learning and Memory Assessment

To assess the deficits in verbal learning induced by THC, we used the Rey Auditory Verbal Learning Test (RAVLT), which has been shown to be sensitive to THC effects in previous studies (Ranganathan et al. 2017). The RAVLT is a 15-word list learning task of verbal memory and hippocampal function. It includes five alternate versions and encompasses five learning trials, an interference list, and both free short- and long-delayed recall and recognition recall trials (Schmidt 1996). To minimize practice effects, we used alternative RAVLT forms and a counterbalanced design to address any differences in form difficulty. The RAVLT was administered once during the test day.

### 2.4 Study Drug

This study used oral THC in the form of dronabinol (10 mg capsule) which is approved by the FDA to treat anorexia in AIDS and other wasting diseases, and emesis in cancer patients undergoing chemotherapy, dronabinol is a synthetic form of THC. When consumed orally, THC exhibits a distinct pharmacokinetic profile compared to when inhaled. Specifically, oral THC shows slower absorption, lower and more delayed peak concentrations, and its effects manifest slower (between 30 to 120 minutes post-consumption) but persist longer (Huestis 2007; Lemberger et al. 1971; Ohlsson et al. 1980; Reyes et al. 1973; Wall et al. 1983). The placebo capsules were identical to dronabinol in shape, color, size, and taste. The order of drug assignment for individuals was determined using computer-generated block randomization and was counterbalanced by the research pharmacist. On each test day, the unblinded research pharmacist prepared the appropriate capsule and provided it to the blinded clinician who administered the drug.

### 2.5 Test Session

Participants were required to arrive at the test facility at 8 AM after fasting overnight. Participants were not permitted to drive to and from the test facility, and transportation was provided if needed. On the morning of each test day, urine drug and pregnancy tests (in women) were conducted to rule out pregnancy and any recent substance use. Weight, height, and body mass index were calculated for all subjects. IV lines were inserted, and baseline behavioral, subjective, and physiological measures were collected. Participants were provided with a standard light breakfast 90 minutes before THC administration. After the administration of dronabinol, outcome measures were repeated periodically throughout the test day as described below, except for the verbal learning and memory outcomes (RAVLT), which were assessed once, 80 minutes after the drug administration. Vital signs (heart rate) were assessed as part of the medical monitoring of the subjects. **Figure 1** represents an approximate schedule of testing. Test days were at least three days apart consistent with data demonstrating undetectable THC levels 48 hours after administration of a single dose of dronabinol (Parikh et al. 2016).

### 2.6 Statistical analyses

Initially, data were examined descriptively using means, standard deviations, and graphs. Each outcome was tested for normality using Kolmogorov–Smirnov test statistics and normal probability plots. Subjective (VAS) and psychomimetic and dissociative (CADSS, PSI) effects were analyzed as peak changes from baseline. These along with verbal learning and memory (RAVLT) outcomes were highly skewed, even after log transformation. Thus, these outcomes were analyzed using the nonparametric approach for repeated measures data by Brunner et al. (2002), in which data first are ranked and then fitted using a mixed-effects model with an unstructured variance-covariance matrix and p values adjusted for analysis of variance-type statistics (ATS). In the models, sex was included as a between-subjects factor and THC (active, placebo) was included as a within-subjects factor. The THC by sex interaction was modeled with subject as the clustering factor. Physiological outcomes (e.g., heart rate) were sufficiently normal and analyzed as peak changes from baseline using linear mixed models with the same factors as above. In terms of power (Heinze 2008), assuming a two-tailed alpha threshold of 0.05, the study sample size of n=56 (33 women, 23 men) provides 80% statistical power to detect between-subject effects of d=0.63 (Mann-Whitney test) and within-subjects effects of d’=0.41 (Wilcoxon test) for females and d’=0.49 for males. Data were analyzed using SAS, version 9.4 (SAS Institute, Cary, NC).

## 3. Results

### 3.1 Participants

Of the 56 participants who completed at least one test day, 23 were men and 33 were women **(Figure S1)**. Men and women showed no significant differences in baseline characteristics such as age, education, race, ethnicity, and BMI (**Table 1**). Both groups had comparable prior exposure to cannabis, and the majority of participants had used cannabis less than five times in the month leading up to the screening (**Table 2**).

**Table 1:** Baseline characteristics of participants.

| Participant characteristics |  |  |  |  |  |
| --- | --- | --- | --- | --- | --- |
|  | Men |  | Women |  | P value |
|  | Mean/n | SD | Mean/n | SD |  |
| Total # (n) | 23 |  | 33 |  |  |
| # on HCM<sup>1</sup> |  |  | 19 (57.6%) |  |  |
| Age (years) | 29.0 | 5.8 | 26.2 | 5.4 | 0.07 |
| Education (years) | 15.6 | 1.3 | 15.4 | 2.2 | 0.65 |
| Race (n) |  |  |  |  | 0.42 |
| Asian | 0 (0%) |  | 3 (9.1%) |  |  |
| Black | 4 (17.4%) |  | 7 (21.2%) |  |  |
| Caucasian | 17 (73.9%) |  | 19 (57.6%) |  |  |
| Mixed | 1 (4.3%) |  | 2 (6.1%) |  |  |
| Other | 0 |  | 0 |  |  |
| Ethnicity |  |  |  |  | 0.99 |
| Hispanic | 6 (26.1%) |  | 9 (27.2%) |  |  |
| Non-Hispanic | 17 (73.9%) |  | 23 (69.7%) |  |  |
| BMI | 25.3 | 3.7 | 25.3 | 4.5 | 0.95 |
| Height (ins) | 70.0 | 3.1 | 63.6 | 3.2 | >0.01 |
| Weight (lbs) | 176.9 | 31.4 | 148.3 | 33.6 | >0.01 |

Abbreviation: BMI, Body Mass Index; HCM, Hormonal Contraceptive medication.
<sup>1</sup> For women who were not using HCM, all the test days were conducted in the early follicular phase. The details of HCMs can be found in **Table S2**.

**Table 2:** Cannabis use history in participants at baseline.

| **Cannabis use history** |  |  |  |
| --- | --- | --- | --- |
|  | **Men** | **Women** | **P value** |
| # of participants | **23** | **33** |  |
| Age on first cannabis exposure | **Mean (SD)**<br>17.7 (3.5) | **Mean (SD)**<br>17.7 (3.7) | 0.98 |
| Last month's cannabis exposure # of days | **N (%)** | **N (%)** | 0.95 |
| 0 days | 9 (53%) | 10 (45%) |  |
| 1-5 days | 6 (35%) | 9 (41%) |  |
| 6-10 days | 1 (6%) | 1 (4%) |  |
| >10 days | 1 (6%) | 2 (9%) |  |
| Time since Last exposure to cannabis | **N (%)** | **N (%)** | 0.40 |
| <1 day | 0 (0%) | 1 (5%) |  |
| 1 day to <1 week | 2 (11%) | 5 (24%) |  |
| 1 week to <1 month | 6 (33%) | 4 (19%) |  |
| 1 month to <6 month | 4 (22%) | 8 (38%) |  |
| 6 month to 1 year | 1 (6%) | 1 (5%) |  |
| > 1 year | 5 (28%) | 2 (9%) |  |
| Lifetime times of cannabis use | **Mean (SD)**<br>1571 (3676) | **Mean (SD)**<br>318.5 (615.4) | 0.18 |
| Lifetime grams of cannabis used | **Mean (SD)**<br>401.2 (994.5) | **Mean (SD)**<br>105.3 (213.4) | 0.25 |
| Last month grams of cannabis used | **Mean (SD)**<br>0.5 (0.9) | **Mean (SD)**<br>0.4 (0.9) | 0.87 |

### 3.2 Subjective Psychoactive Effects (VAS “high”)

A significant THC by sex interaction effect was observed (ATS=3.81, num df=1, p=0.05, Cohen’s d=0.43) such that a significant main effect of dose (THC) was observed among women (ATS=4.76, num df=1, p=0.03, Cohen’s d’=0.48) but not men (ATS=0.49, num df=1, p=0.48, d’=0.11). The effects are depicted in **Figure 2** and tabulated in **Tables S3 and S4**.

**Figure 2:**
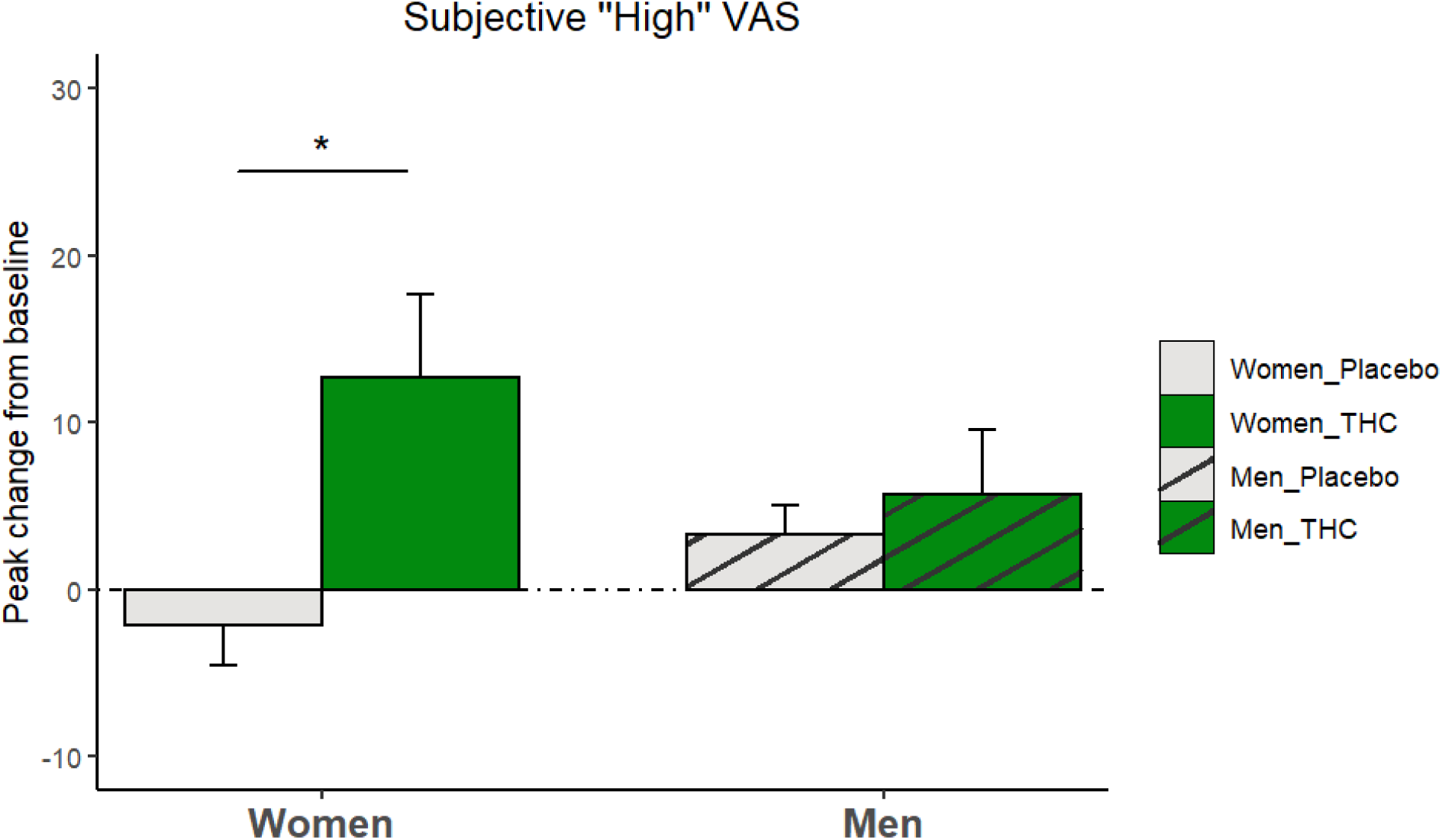
The mean and standard error of peak changes in visual analog scale (VAS) of subjective “high” perception from baseline categorized by THC/placebo administration and sex. The VAS was scored from 0 to 100. The Asterisk indicates statistically significant differences.

### 3.3 Psychotomimetic and Dissociative Effects

#### CADSS

There was a main effect of dose (THC) in both observer-rated (ATS= 31.3, num df=1, p=0.0001) and subjective-items (dose effect: ATS= 42.0, num df=1, p=0.0001) CADSS scores compared to placebo. No main effects of sex in observer-rated (ATS= 0.47, num df=1, p= 0.49) or subjective-items scores (ATS: 2.77, num df=1, p=0.9) were observed. Additionally, no dose by sex interaction effects in observer-rated (ATS= 0.19, num df=1, p=0.66) or subjective-rated items score (ATS=2.21, num df=1, p=0.14) was observed.

#### PSI

There was a main effect of dose (THC) on psychotomimetic effects measured using PSI scores (ATS= 52.7, num df=1, p<0.0001). There was no main effect of sex (ATS=0.46, num df=1, p=0.50) or THC by sex interaction effect (ATS=0.1, num df=1, p=0.76) (**Figure 3 and Table S4**).

**Figure 3:**
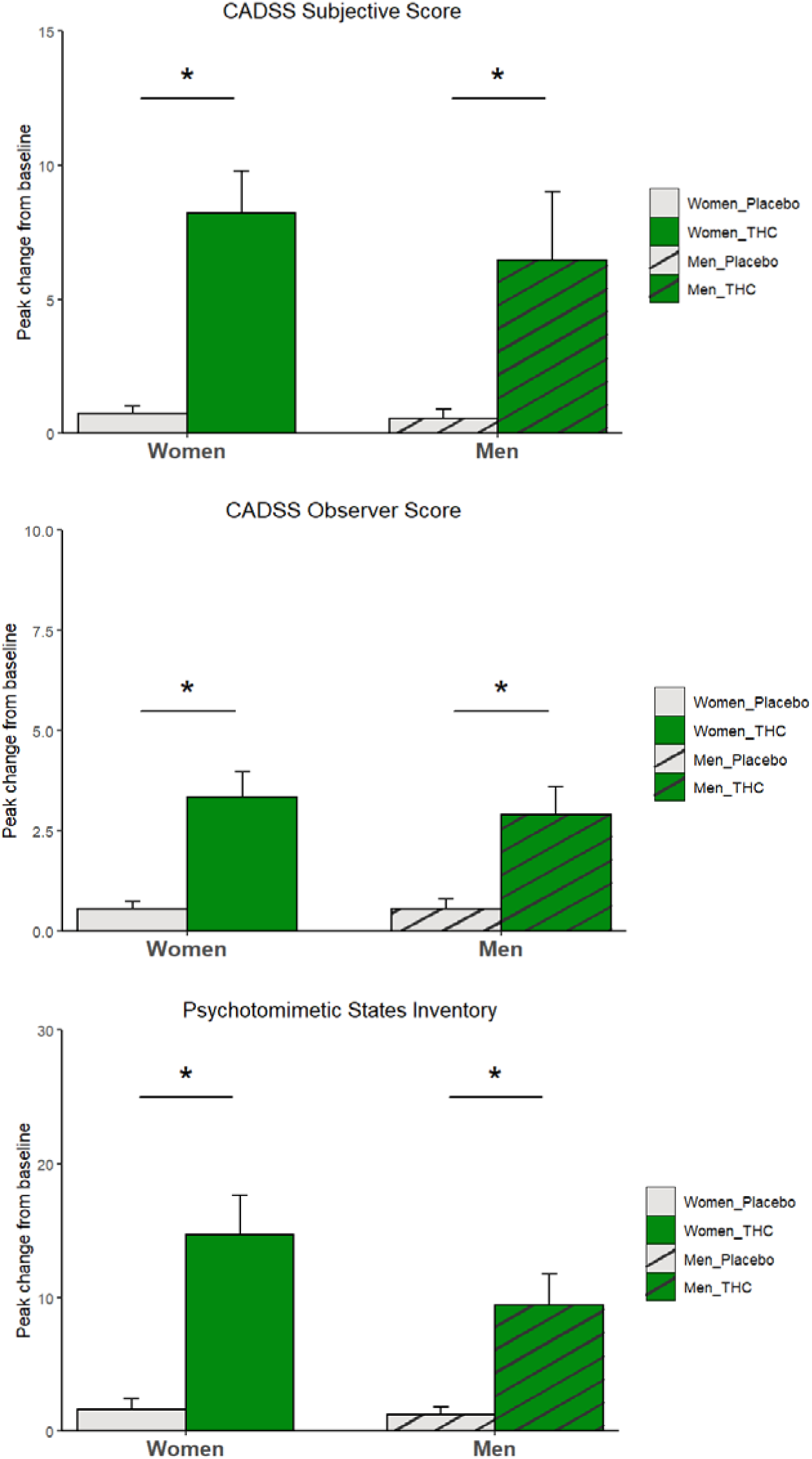
The mean and standard error of peak changes in psychotomimetic scales scores from baseline categorized by THC/placebo administration and sex. Clinician-administered dissociative states scale (CADSS) subjective items score (top), observer-rated score (middle), and psychotomimetic states inventory score (bottom). Asterisks indicate statistically significant differences.

### 3.4 Verbal Learning and Memory

The mean, standard deviation, and regression model results of the various RAVLT outcomes are tabulated in **Table S5**. There was no main effect of dose (THC), sex, or dose by sex interaction in the immediate, short-delay, or long-delay recall.

### 3.5 Physiological Effect

Significant increases in heart rate were observed following THC administration (dose effect: F (1, 54) =21.63, p<0.0001). However, there was no main effect of sex (F (1, 54) =1.53, p= 0.22) or THC by sex interaction (F (1, 54) =1.52, p= 0.22) on heart rate (**Figure 4**).

**Figure 4:**
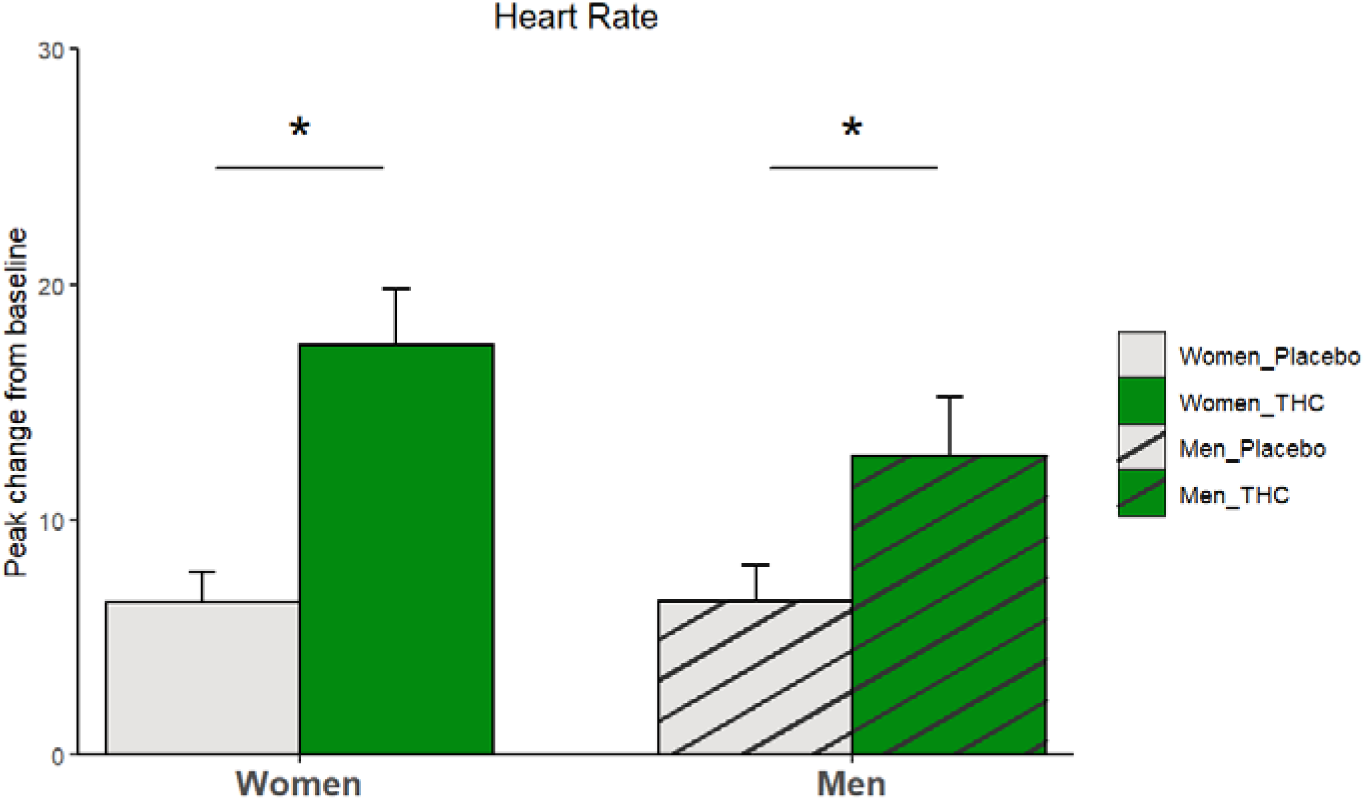
The mean and standard error of participants’ peak change in heart rate from baseline categorized by THC/placebo administration and sex. Asterisks indicate statistically significant differences.

## 4. Discussion

In this double-blind, placebo-controlled, randomized human laboratory study, we examined the subjective psychoactive, psychotomimetic, and physiological effects and verbal learning and memory deficits following a single dose of 10 mg oral THC in healthy men and women without CUD. We observed a significantly greater THC-induced “high” among women compared to men. This differential subjective experience was selective to THC-induced “high” as men and women experienced equivalent psychotomimetic, physiological, and cognitive effects after THC administration. Indeed, the lack of verbal memory deficits at this dose, suggests that women experienced discernable subjective effects even at a dose of THC that failed to produce cognitive effects. This study was not designed to examine the abuse liability of THC. However, the observed pattern is consistent with others suggesting a greater selective vulnerability to some effects of THC amongst women. Whether this pattern is supportive of a heightened addictive potential among women compared to men, needs to be further elucidated.

Although the magnitude of difference in subjective “high” between women and men was small (effect size: d=0.43), the directionality is consistent with previous studies of IV and sublingual THC, and smoked cannabis (Bassir Nia et al. 2022; Cooper and Haney 2009; Lake et al. 2023; Makela et al. 2006). In addition, Sholler et al. (2021) also observed greater subjective effects among women, although some of the effects such as anxiety and restlessness may be unpleasant. However, our findings are not consistent with Penetar et al. (2005) and Haney (2007) who found that men were more sensitive to the subjective effects of smoked cannabis and oral THC, respectively. While the majority of the published literature to date suggests that women are more sensitive to the acute subjective effects of THC, there are some mixed results that need further study to address several methodological differences that may underlie the results-namely, dose, route, and formulation of THC used and participant characteristics including cannabis use frequency and menstrual cycle variability in women. Our study included women on hormonal contraception or only in the early follicular phase to reduce the differential impact of ovarian steroids across the menstrual cycle. Also, we included only healthy individuals without CUD who had minimal cannabis exposure in the past month. Studying individuals with a broader range of recent cannabis exposure may help with greater generalizability of the results given the currently increasing rates of cannabis use. Our study included only a single dose of oral THC (10mg). Future studies should also examine a wide dose range as the sex differences may vary with increasing doses of THC (Bassir Nia et al. 2022; Fogel et al. 2017).

Several factors may underlie the observed differences in our study including sex differences in THC’s distribution and metabolism. Lunn et al. (2019) have shown that when fasting, women exhibit notably higher plasma THC concentrations after ingestion of a 5 mg oral THC dose. Similarly, Spindle et al. (2020) found that women have higher THC and THC metabolites after ingestion of brownies containing cannabis, a difference that authors suggested might not be only due to differences in BMI. Furthermore, oral THC undergoes first-pass metabolism in the liver, producing active metabolites like 11-OH-THC, which might be influenced by sex-specific hepatic metabolism (Lunn et al. 2019; Nadulski et al. 2005; Narimatsu et al. 1991; Wall et al. 1983). It is important to note that we have previously reported similar sex differences in subjective effects using IV THC that avoids the confounds of first-pass metabolism (Bassir Nia et al. 2022) suggesting that the observed pattern cannot be fully explained by sex differences in hepatic metabolism.

It is noteworthy, that our results are consistent with the robust preclinical literature demonstrating sex differences in THC effects that may be modulated by gonadal steroids (Bassir Nia et al. 2018; Calakos et al. 2017; Cooper and Craft 2018). Compared to male rats, female rats showed increased sensitivity to THC-reinforcing effects, as evidenced by their faster acquisition of THC self-administration behaviors (Fattore et al. 2007; Freels et al. 2023) and enhanced cue- and drug-induced reinstatement of THC-seeking behaviors (Fattore et al. 2010).

Interestingly, the sensitivity to the reinforcing effects of THC showed a reduction in gonadectomized female rats (Fattore et al. 2007; Fattore et al. 2010). Marusich et al. (2015) further support a role for sex hormones as gonadectomized female rats showed higher withdrawal signs of THC when supplied with estradiol and progesterone but on the other hand, female gonadectomized rats showed less withdrawal signs of THC when supplied with testosterone. Following THC exposure, female rats show higher downregulation or desensitization of CB1-R across all brain regions (Burston et al. 2010; Farquhar et al. 2019). Likewise, the CB1-R selective antagonist rimonabant was up to 10 times more potent in female rats than male rats in blocking the antinociceptive effects of THC (Craft et al. 2012). Preclinical data has demonstrated that cannabinoid drugs (i.e., both cannabinoid agonists and antagonists) bind with greater affinity to CB1-Rs in female rats compared to male rats (Craft et al. 2012), which may also explain the differential effects of cannabinoid agonists in women as observed in our studies. Importantly, there are well-documented sex differences in the endocannabinoid system demonstrating sex-related regional variations in CB1-R availability (Farquhar et al. 2019; Liu et al. 2020; Llorente-Berzal et al. 2013; Paola Castelli et al. 2014), that may be influenced by gonadal hormones (De Fonseca et al. 1994). Some human brain imaging studies have compared CB1-Rs availability in men and women with mixed results (Laurikainen et al. 2019; Neumeister et al. 2013; Normandin et al. 2015; Radhakrishnan et al. 2022; Van Laere et al. 2008). Further human studies, accounting for the differences in radioligands and the influence of gonadal hormones are needed to fully understand the sex differences in the endocannabinoid system in humans.

While our study provides valuable insights, some limitations warrant further discussion. Oral THC is associated with inter and intra-individual variability influenced by body fat percentage and liver-based phase I THC metabolism that was not assessed in this study. Further, dronabinol is a commercially available, synthetic form of THC, the acute effects of which may differ from commonly used edible cannabis products. We selected dronabinol to ensure uniform and fixed dosing across participants to avoid THC dose related variability in effects. Similarly, the findings from this study need to be interpreted keeping the dose and route of THC in mind and cannot be extrapolated to vaporized or smoked cannabis. Our study employed a single low dose that produced some subjective and physiological effects but no verbal learning deficits in both men and women. Future studies should examine a wider dose range as cannabis-related sex differences may vary in different doses and include individuals with a wide range of cannabis exposures.

## 5. Conclusion

We report selective sex differences in some (psychoactive subjective effect of “high”) but not other (psychotomimetic, physiological effects or verbal learning deficits) THC-induced effects in healthy individuals without CUD such that women experienced greater self-reported THC-induced “high” compared to men. These findings are consistent with the preclinical literature and preponderance of clinical data thus far and stress the importance of sex-specific approaches to evaluation, prevention, and treatment strategies, as women represent a growing segment of people using THC-based products.

## Data Availability

All data produced in the present study are available upon reasonable request to the authors

## Notes

**Funding** This study was funded by the National Institute on Drug Abuse (NIDA); grant number 1R21DA038821-01A1 (P.I MR) and VA Connecticut Healthcare System (VACHS). J.P.D. is supported by the grants K23DA052682 and R21DA057240 from the National Institute of Drug Abuse (NIDA). ABN is supported by the National Institute of Health (NIMH) K12DA000167 grant.

**Conflict of interest statement** J.P.D. has been supported in clinical trials by Jazz Pharmaceuticals, specifically through medication provisions. Additionally, J.P.D. has been a compensated consultant for Boehringer Ingelheim. A.B.N is a member of the Scientific Advisory Committee of Synendos Therapeutics AG, Switzerland.

### Competing Interest Statement

J.P.D. has been supported in clinical trials by Jazz Pharmaceuticals, specifically through medication provisions. Additionally, J.P.D. has been a compensated consultant for Boehringer Ingelheim. A.B.N is a member of the Scientific Advisory Committee of Synendos Therapeutics AG, Switzerland.

### Clinical Trial

NCT02781519

### Funding Statement

This study was funded by National Institute on Drug Abuse (NIDA); grant number 1R21DA038821-01A1. J.P.D. is supported by the grants K23DA052682 and R21DA057240 from the National Institute of Drug Abuse (NIDA). ABN is supported by the National Institute of Health (NIMH) K12DA000167 grant.

### Author Declarations

The study protocol was approved by the Institutional Review Boards of the VA Connecticut Healthcare System and Yale University School of Medicine.

### Summary of Updates

Details about the design have been added. The discussion section has been shortened. The analysis of the heart rate data has been changed to peak-from-baseline to be consistent with other outcomes. Complementary data has been added to the supplementary material.

