## supplementary material for "Sex Differences in the Acute Effects of Oral THC: A Randomized, Placebo-Controlled, Crossover Human Laboratory Study"

Table S1: A brief review of the previous human laboratory studies investigating the sex differences in acute effects of oral THC-containing products

| Previous studies investigating oral THC-containing products | | | | | |
| --- | --- | --- | --- | --- | --- |
| Reference | **Design** | **Population** | **Intervention/ Comparison** | **Outcome** | **Result** |
| Haney (2007) | Randomized double-blind cross-over study | Substudy 1: Current frequent cannabis smokers  Substudy 2: No history of frequent cannabis use | Substudy 1: Oral THC (Marinol) 20 and 40 mg and placebo  Substudy 2: Oral THC (Marinol) 2.5 and 5 and 10 mg and placebo  In both substudies, the THC was administered 30 minutes after oral naltrexone (12 mg) or placebo | Acute psychoactive effect  Blood pressure  Heart rate  Performance battery^1^  Immediate/delayed recall tests | Substudy 1: no sex differences in any of the outcomes  Substudy 2: In placebo pretreatment condition, men reported higher subjective psychoactive effect (statistical significance not reported) |
| Fogel (2017) | Pooled data from 7 Randomized double-blind cross-over studies | Current cannabis users | Oral THC (dronabinol) 5, 15, a high dose of 25 or 30, and placebo | Acute psychoactive effect  Heart rate  Blood pressure  Temperature  Performance battery^2^ | Women reported higher psychoactive effects on 5 mg dose trials, but men reported higher psychoactive effects on 15 mg dose trials  No sex differences were observed in other variables |
| Scholler (2020) | Pooled data from 4 randomized double-blind studies (3 cross-over and 1 between-subject studies) | Infrequent cannabis users (no use in the 30 days preceding randomization) | Oral cannabis containing low dose (5 or 10 mg) and high dose (20 or 25 mg) THC and placebo | Acure psychoactive effect  Heart rate  Blood pressure  Cognitive battery^3^  Plasma concentration of THC, 11-OH-THC, and THC-COOH | The acute psychoactive feeling of “drug effect” was higher in women following low-dose THC and “restless” and “anxious/nervous” following high-dose THC  No sex differences were observed in other variables |
| MacNair (2023) | Pooled data from 2 randomized double-blind between-subject studies | Healthy individuals | THC-containing oral cannabis product with low-dose (~2.5 or ~5 mg THC) or high-dose (~7.5 or ~10mg THC) or placebo twice daily for 7 days | Psychoactive effects  Adverse events | Women reported greater subjective “relaxed” effects and more adverse events in the high-dose group. |

^1^ Included a three-minute digit-symbol substitution task, a three-minute repeated acquisition task, a ten-minute divided attention task, a ten-minute rapid information task, and immediate and delayed digit-recall tasks.

^2^ Including the repeated acquisition of response sequences task, digit-symbol-substitution test, and time reproduction task.

^3^ Including digit symbol substitution task, paced serial addition task, and divided attention task.

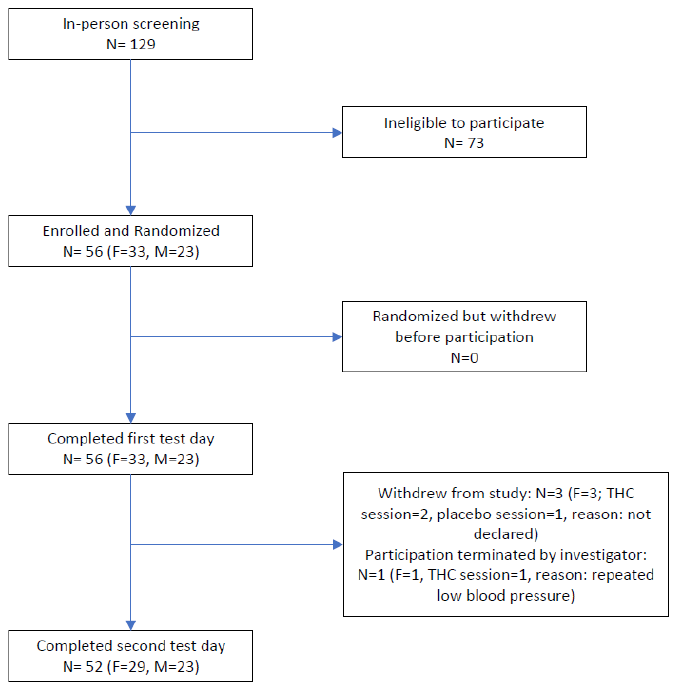

Figure S1: The CONSORT flow diagram of participants' selection and enrolment

Table S2: The details of contraceptive medication in women participants who used hormonal contraceptive medication.

| **Hormonal contraceptive medication** | | | |
| --- | --- | --- | --- |
| **# of participant** | **Route of medication** | **Medication and dosage** | **Duration of using current medication (approximate)** |
| 1 | IUD | levonorgestrel; 52 mg  (20 mcg/day release) | 11 months |
| 2 | IUD | levonorgestrel; 52 mg  (20 mcg/day release) | 3 years and 11 months |
| 3 | IUD | levonorgestrel; 52 mg  (20 mcg/day release) | 6 years and 11 months |
| 4 | Oral | Norethindrone acetate; 1mg  Ethinyl estradiol; 0.02 mg | 2 years |
| 5 | IUD | 13.5 mg of Levonorgestrel; 13.5 mg  (14 mcg/day release) | 1 year |
| 6 | IUD | Levonorgestrel; 19.5 mg  (17.5 mcg/day release) | 4 months |
| 7 | Oral | Norethindrone acetate, N/A Ethinyl estradiol, N/A | 3 years |
| 8 | Implant | Etonogestrel; 68 mg  (60-70 mcg/day release) | 4 months |
| 9 | Oral | Norgestimate, 0.25 mg  Ethinyl estradiol, 35 mcg | 5 years |
| 10 | Oral | Norethindrone acetate, N/A Ethinyl estradiol, N/A | 1 year |
| 11 | Oral | Levonorgestrel 0.1mg,  Ethinyl Estradiol 0.02mg | 5 years |
| 12 | Oral | Levonorgestrel, 0.15 mg  Ethinyl estradiol, 30 mcg | 9 months |
| 13 | Patch | Norelgestromin, 150 mcg daily release  Ethinyl estradiol, 35 mcg daily release | 11 months |
| 14 | Vaginal ring | Etonogestrel, 0.12 mg daily release  Ethinyl estradiol, 0.015 mg daily release | 1 year and 6 months |
| 15 | Oral | Norgestimate 0.18-0.25 mg, Ethinyl estradiol, 35mcg | 2 months |
| 16 | Implant | Etonogestrel, 68 mg | 2 years |

Abbreviation: IUD, Intrauterine Device; N/A, Not Available.

The data on the type and dosage of hormonal contraception was not available for three women using these medications.

Table S3: The timing of peak "high" psychoactive effect following THC administration

| **Timing^1^ of peak-change-from-baseline of “high” psychoactive effect during the THC test days** | | | | |
| --- | --- | --- | --- | --- |
|  | **# of participants (%)** | | | |
|  | **80 minutes** | **150 minutes** | **180 minutes** | **300 minutes** |
| Women (N=32) | 13 (39.5%) | 7 (21.2%) | 8 (24.2%) | 4 (12.1%) |
| Men (N=23) | 13 (56.5%) | 6 (26.1%) | 2 (8.7%) | 2 (8.7%) |

^1^Time of Peak subjective effect was defined as the first time-point in which the participants reported the highest magnitude of “high” effects they experienced on the visual analog scale after receiving THC.

Table S4: The mean and standard deviation of various acute psychoactive effects and psychotomimetic effects of THC at the baseline and at peak-change-from-baseline

| **Peak acute psychoactive and psychotomimetic effects** | | | | | | | | | | |
| --- | --- | --- | --- | --- | --- | --- | --- | --- | --- | --- |
|  |  | | **Baseline** | | | | **Peak minus baseline** | | | |
|  |  | | **Women** | | **Men** | | **Women** | | **Men** | |
|  |  | | **Mean** | **SD** | **Mean** | **SD** | **Mean** | **SD** | **Mean** | **SD** |
| **Visual analog scale of psychoactive states** | **In an altered state** | Placebo | 36.7 | 20.3 | 35.8 | 27.0 | 17.5 | 15.8 | 7.1 | 14.7 |
|  |  | THC | 40.7 | 26.4 | 42.0 | 26.4 | 8.5 | 19.7 | 12.5 | 27.0 |
|  | **Sad** | Placebo | 50.6 | 25.7 | 59.2 | 23.5 | 7.7 | 12.1 | 1.8 | 14.4 |
|  |  | THC | 52.7 | 28.7 | 60.6 | 21.2 | 6.1 | 16.9 | 5.6 | 15.0 |
|  | **Tired** | Placebo | 32.5 | 23.9 | 21.0 | 19.3 | 12.0 | 17.7 | 16.2 | 20.6 |
|  |  | THC | 23.3 | 25.6 | 27.3 | 24.7 | 31.7 | 31.2 | 15.7 | 21.9 |
|  | **Anxious** | Placebo | 13.1 | 19.3 | 9.8 | 16.0 | -1.1 | 16.8 | -0.1 | 10.1 |
|  |  | THC | 10.0 | 15.5 | 7.1 | 8.9 | 9.2 | 26.9 | 9.0 | 21.8 |
|  | **Suspicious** | Placebo | 9.0 | 16.5 | 6.8 | 14.3 | -3.3 | 13.4 | -1.2 | 6.3 |
|  |  | THC | 3.6 | 7.6 | 4.4 | 5.4 | 3.4 | 9.1 | -0.3 | 5.5 |
|  | **Calm** | Placebo | 60.6 | 24.2 | 64.0 | 26.2 | 12.8 | 17.4 | 0.7 | 12.4 |
|  |  | THC | 60.6 | 25.5 | 60.5 | 26.1 | 10.8 | 17.5 | 9.5 | 21.4 |
|  | **Happy** | Placebo | 4.6 | 9.0 | 2.8 | 4.1 | -1.0 | 6.1 | 0.5 | 3.1 |
|  |  | THC | 2.2 | 5.0 | 2.7 | 3.6 | 0.9 | 4.1 | 3.0 | 10.2 |
|  | **High** | Placebo | 12.1 | 18.8 | 6.3 | 9.0 | -2.2 | 12.6 | 3.3 | 8.5 |
|  |  | THC | 9.9 | 14.8 | 5.8 | 8.2 | 12.7 | 28.1 | 5.7 | 18.3 |
|  | **Puzzled** | Placebo | 31.1 | 23.0 | 30.9 | 26.5 | 10.3 | 12.1 | 3.4 | 14.4 |
|  |  | THC | 32,0 | 22.6 | 27.3 | 23.3 | 7.3 | 16.9 | 14.3 | 15.0 |
|  | **Irritable** | Placebo | 6.8 | 11.8 | 3.5 | 7.1 | -2.8 | 6.3 | -0.2 | 4.0 |
|  |  | THC | 3.2 | 5.3 | 2.6 | 3.8 | 5.5 | 14.0 | 2.6 | 8.2 |
|  | **Hungry** | Placebo | 48.1 | 27.7 | 42.9 | 33.3 | 7.7 | 15.1 | 1.8 | 18.2 |
|  |  | THC | 51.3 | 29.7 | 40.0 | 31.3 | 6.1 | 20.5 | 5.6 | 24.5 |
|  | **Strong** | Placebo | 2.1 | 4.4 | 1.6 | 2.6 | 10.8 | 20.3 | 3.2 | 6.2 |
|  |  | THC | 1.3 | 2.8 | 1.2 | 1.3 | 57.8 | 30.2 | 26.8 | 27.5 |
|  | **Clear in thoughts** | Placebo | 6.2 | 19.2 | 2.3 | 3.8 | 7.7 | 12.1 | 1.8 | 14.4 |
|  |  | THC | 2.1 | 4.7 | 1.7 | 2.8 | 6.1 | 16.9 | 5.6 | 15.0 |
|  | **Worried** | Placebo | 1.5 | 3.2 | 1.7 | 2.6 | -0.1 | 3.0 | 0.9 | 2.4 |
|  |  | THC | 1.0 | 2.7 | 1.7 | 2.8 | 4.3 | 12.2 | 1.7 | 4.6 |
|  | **Physically well** | Placebo | 5.4 | 9.5 | 3.8 | 4.9 | 7.7 | 22.0 | 1.8 | 4.3 |
|  |  | THC | 5.3 | 10.7 | 3.7 | 10.2 | 6.1 | 13.2 | 5.6 | 17.8 |
|  | **Safe** | Placebo | 43.3 | 26.8 | 26.6 | 23.6 | 8.4 | 18.8 | 20.1 | 27.4 |
|  |  | THC | 29.6 | 25.0 | 25.3 | 25.8 | 28.7 | 29.2 | 19.8 | 23.5 |
|  | **Focused** | Placebo | 26.3 | 27.7 | 27.4 | 26.3 | 26.2 | 24.0 | 26.0 | 25.9 |
|  |  | THC | 17.8 | 23.6 | 27.0 | 27.1 | 33.3 | 31.0 | 29.0 | 21.9 |
| **CADSS** | Observer-rated | Placebo | 0.0 | 0.0 | 0.3 | 0.7 | 0.6 | 1.0 | 0.6 | 1.1 |
|  |  | THC | 0.0 | 0.0 | 0.0 | 0.2 | 3.3 | 3.6 | 2.9 | 3.4 |
|  | Subjective | Placebo | 0.1 | 0.3 | 0.5 | 0.9 | 0.7 | 1.6 | 0.6 | 1.6 |
|  |  | THC | 0.0 | 0.0 | 0.2 | 0.6 | 8.2 | 8.8 | 6.5 | 12.2 |
| **PSI** | Total score | Placebo | 9.2 | 6.1 | 9.5 | 3.9 | 1.6 | 4.3 | 1.2 | 2.7 |
|  |  | THC | 8.5 | 2.8 | 9.0 | 2.6 | 14.7 | 16.8 | 9.4 | 11.0 |

Abbreviations: CADSS, Clinician-Administered Dissociative Symptoms Scale; PSI, Psychotomimetic States Inventory.

Table S5: The results of regression models of Rey Auditory Verbal Learning Test (RAVLT) outcomes

| **RAVLT outcomes** | | | | | | | |
| --- | --- | --- | --- | --- | --- | --- | --- |
|  | **Women [mean (SD)]** | | **Men [mean (SD)]** | | **Linear regression model results** | | |
|  | **THC** | **Placebo** | **THC** | **Placebo** | **Effect** | **Estimate (ATS)** | **P value** |
| Total immediate recall | 51.4 (10.5) | 52.7 (11.1) | 52.1 (12.8) | 51.6 (13.5) | Dose | 1.22 | 0.27 |
|  |  |  |  |  | Sex | 0.02 | 0.89 |
|  |  |  |  |  | Dose x Sex | 0.56 | 0.45 |
| Short delay recall | 11.1 (3.1) | 11.0 (3.3) | 11.8 (3.2) | 11.7 (3.8) | Dose | 0.06 | 0.80 |
|  |  |  |  |  | Sex | 1.64 | 0.20 |
|  |  |  |  |  | Dose x Sex | 0.04 | 0.84 |
| Long delay recall | 10.7 (3.1) | 10.3 (3.4) | 10.8 (3.5) | 11.2 (3.3) | Dose | 0.1 | 0.75 |
|  |  |  |  |  | Sex | 0.27 | 0.60 |
|  |  |  |  |  | Dose x Sex | 0.63 | 0.43 |
| Short recall minus T5 | -1.5 (1.9) | -1.9 (2.2) | -0.6 (2.1) | -1.0 (2.6) | Dose | 0.21 | 0.65 |
|  |  |  |  |  | Sex | 7.24 | **0.007** |
|  |  |  |  |  | Dose x Sex | 0.13 | 0.71 |
| Total Intrusions | 3.7 (4.0) | 2.4 (3.2) | 2.6 (3.0) | 4.4 (5.6) | Dose | 0.52 | 0.48 |
|  |  |  |  |  | Sex | 0.03 | 0.87 |
|  |  |  |  |  | Dose x Sex | 2.2 | 0.14 |
| Total perseveration | 3.8 (5.1) | 3.2 (3.7) | 2.6 (2.7) | 5.0 (5.5) | Dose | 3.27 | 0.07 |
|  |  |  |  |  | Sex | 0 | 0.94 |
|  |  |  |  |  | Dose x Sex | 1.4 | 0.24 |
| Interference | 6.0 (2.0) | 6.5 (2.2) | 6.6 (2.5) | 6.0 (2.6) | Dose | 0.19 | 0.67 |
|  |  |  |  |  | Sex | 0.06 | 0.81 |
|  |  |  |  |  | Dose x Sex | 3.15 | 0.08 |

Abbreviations: RAVLT, Rey Auditory Verbal Learning Test.
